# Kinetics of SARS-CoV-2 Antibody Avidity Maturation and Association with Disease Severity

**DOI:** 10.1101/2020.07.30.20165522

**Authors:** Yiqi Ruben Luo, Indrani Chakraborty, Cassandra Yun, Alan H.B. Wu, Kara L. Lynch

## Abstract

The kinetics of IgG avidity maturation during SARS-CoV-2 infection was studied. The IgG avidity assay used a novel label-free immunoassay technology. It was found that there was a strong correlation between IgG avidity and days since symptom onset, and peak readings were significantly higher in severe than mild disease cases.

## Main Text

In the race to identify correlates of COVID-19 immunity, the focus has been on characterizing the production of SARS-CoV-2 antibodies. Most immunocompetent individuals with symptomatic infections develop detectable SARS-CoV-2 antibodies within 2 weeks of symptom onset.^1^ The SARS-CoV-2 IgG antibody response is more robust in severe cases of COVID-19 at all time-points after seroconversion.^1^ Preliminary findings suggest that the magnitude of the antibody response correlates with virus neutralizing power, but some hypothesize that the antibodies could promote disease progression.^2^ Influential factors in the distinction between protective antibodies and those that promote pathology include antibody isotype, concentration, specificity, post-translational modifications, and avidity for the virus. Most SARS-CoV-2 antibody studies to date have focused on the quantity of specific isotypes rather than the quality of the antibodies. Antibody avidity, or functional affinity, is a measure of the maturation of the humoral immune response after both infection and vaccination and typically increases over time. For some viruses, the measurement of antibody avidity is used to differentiate acute infection from past exposure or vaccination (e.g. toxoplasmosis, SARS-CoV infection) and to detect re-infection in the presence of prolonged virus-shedding.^3,4^ Here we report the development of a method to characterize SARS-CoV-2 IgG avidity maturation in COVID-19 patients from initial diagnosis through convalescence.

The IgG avidity assay was established on a novel label-free immunoassay platform Gator Analyzer (Gator Bio, Palo Alto, CA) to measure SARS-CoV-2 IgG avidity to the virus spike protein receptor-binding domain (RBD). The platform utilizes thin-film interferometry and can measure the entire time course of formation of an immune complex on a sensing probe without attaching a reporter.^5^ The sensing probe in use was pre-coated with recombinant RBD (Histagged, Sino Biological, Wayne, PA). Serum samples were diluted 10-fold in running buffer (PBS with 0.02% Tween 20, 0.2% BSA, and 0.05% NaN_3_) for analysis. The steps of the IgG avidity assay included 1) dipping the sensing probe in running buffer for a baseline measurement, 2) formation of RBD-IgG immune complex on the sensing probe, 3) dissociation of loosely bound IgG using either running buffer or 3 M urea in running buffer, and 4) formation of RBD-IgG-Anti-IgG immune complex using 10 μg/ml anti-IgG in running buffer (goat anti-human IgG antibody, Jackson Immunoresearch, West Grove, PA). The sensing probe was washed in running buffer for 1 min between the steps, and regenerated in glycine pH 2.0. The signal increase in the final step, which is proportional to the quantity of RBD-IgG-Anti-IgG immune complex on the sensing probe, was measured. As other isotypes of antibodies might bind to RBD in the second step, the measurement of the RBD-IgG-Anti-IgG immune complex enhanced the assay specificity. The IgG avidity index was calculated as the ratio of the readout with the dissociation agent (urea) to the reference (running buffer), presented as a percentage. The performance of the IgG avidity assay was verified for precision and specificity. Precision was evaluated using a spiked serum sample prepared by adding a standard anti-RBD IgG (Absolute Antibody, Oxford, UK) in a negative serum sample. The spiked serum sample was measured simultaneously by a row of 7 sensing probes, and the assay was repeated 4 times with regeneration of the sensing probes. The CV among the sensing probes was 4.5%, and the CV among regeneration cycles was 10.4%. The spiked serum sample was also measured in 4 consecutive days and the CV was 13.7%. Specificity was evaluated using a set of 28 serum samples from individuals tested RT-PCR negative for SARS-CoV-2 and positive for other respiratory viruses (4 coronavirus HKV1, 1 coronavirus 229E, 2 coronavirus OC43, 11 human rhinovirus/enterovirus, 4 human metapneumovirus, 3 respiratory syncytial virus, 2 parainfluenza type 1 virus, 1 adenovirus). No formation of RBD-IgG-Anti-IgG immune complex was observed for these samples, providing negative results in the IgG avidity assay.

The study was approved by the Institutional Review Board of the University of California. All testing was performed on remnant serum samples collected for routine clinical testing and stored at −20°C prior to testing. Individual and serial serum samples (n = 168) from patients with PCR-confirmed COVID-19 (n = 90, 51% male, median age 49, 24% admitted to the ICU) were analyzed. Based on least squares linear regression analysis, there was a strong correlation between IgG avidity and days since symptom onset (P < 0.0001) (Figure 1A). SARS-CoV-2 IgG avidity did not correlate with IgG concentration, as measured previously using a quantitative immunoassay (1). The kinetics of IgG avidity maturation in 13 patients for whom serial samples (≥3) were available is shown in Figure 1B. In all patients, IgG avidity increased over time. Peak measurements, which included the last measurement per 7-day interval per patient, were used to compare IgG avidity by disease severity. Peak readings were significantly higher for specimens from ICU than non-ICU patients for the first month after symptom onset (1-4 weeks) and thereafter, using the t-statistic (P < 0.0001) (Figure 1C).

**Figure 1.**
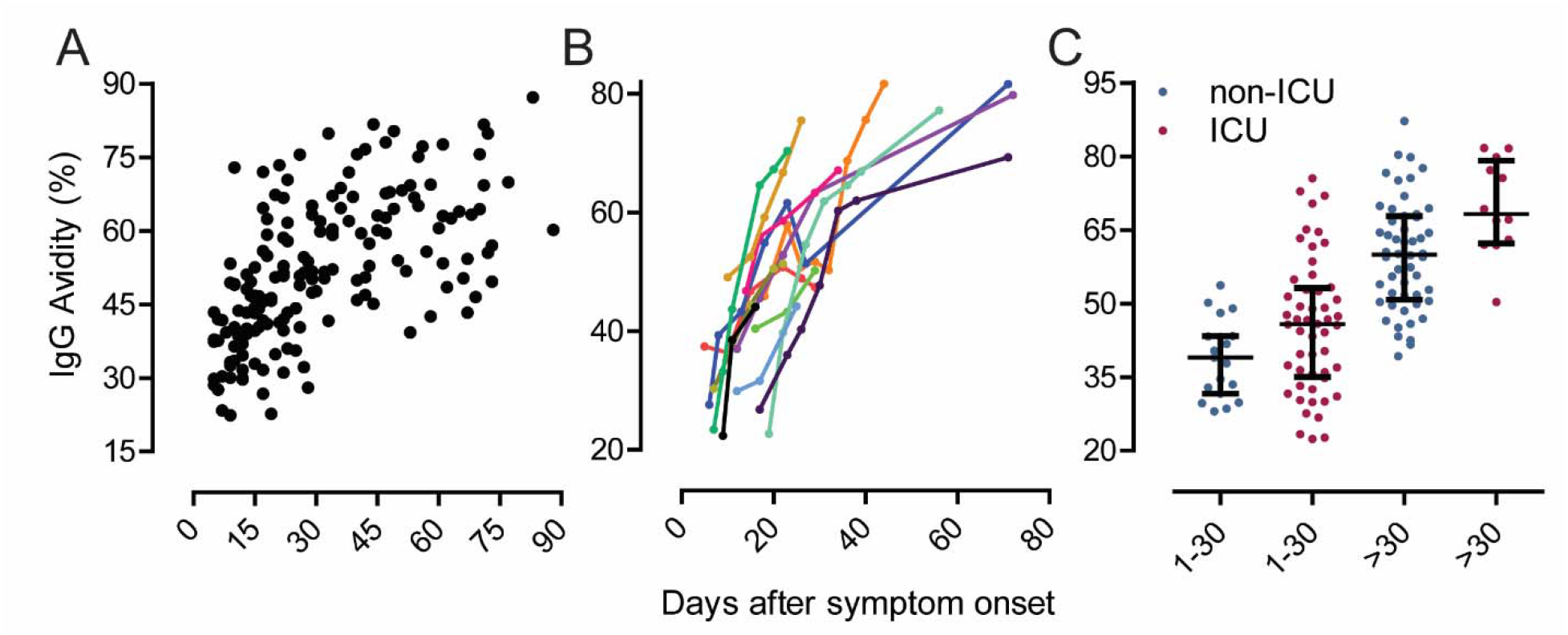
Kinetics of SARS-CoV-2 IgG avidity and distribution of response by level of care (ICU vs non-ICU). (A) Correlation between SARS-CoV-2 IgG avidity and days after symptom onset (n=90 COVID patients, n=168 serum samples). (B) Kinetics of SARS-CoV-2 IgG avidity by days after symptom onset for 13 patients with ≥3 serial samples available for testing. (C) Distribution of IgG avidity by level of care.

To our knowledge this is the first study to evaluate antibody avidity maturation in COVID-19 patients. The findings confirm that IgG avidity significantly increases from 1-90 days post-symptom onset and is significantly associated with disease severity. The findings are consistent for what has been reported for SARS-CoV where avidity indices were low among serum samples collected ≤50 days after symptom onset (30.8% ± 11.6%), intermediate from 51-90 days (52.1% ± 14.1%), and high for samples collected after 90 days (78.1% ± 8.0%).^6^ The availability of samples after 90 days for SARS-CoV-2 in our cohort was limited at the time of this analysis.

For other viruses, the existence of only low-quality and low-concentration antibodies can enhance the infection (antibody-dependent enhancement) and inflammation (antibody-mediated immune enhancement), while high-avidity antibodies capable of blocking receptor binding are typically protective and promote virus neutralization.^7^ Given that SARS-CoV-2 specific IgG avidity is strong in ICU patients after 1 month, these negative effects may not play a role in COVID-19 disease progression and severity. A recent study suggests that the SARS-CoV-2 IgG concentrations can wane over time, however, it is unclear if the antibodies that persist are of high enough quality to neutralize the virus.^8^ Further studies are needed to determine if both the antibody concentration and antibody avidity correlate with virus neutralization and persist over time. The avidity method described here makes use of the label-free immunoassay technology, thin-film interferometry, which allows for direct observation of the time course of immune complex formation. This technology could be a valuable tool for continued characterization of the adaptive immune response to SARS-CoV-2.

## Data Availability

N/A

